# Hormonal Contraceptives Drive Genital Lipid Metabolism Reprogramming and Susceptibility to HIV Infection

**DOI:** 10.64898/2026.06.09.26355168

**Authors:** Abrha G. Gebrehiwot, Maysa Niazi, Yohannes A. Ambaw, Nicola Henriquez, Jocelyn M. Wessels, Julie Lajoie, Joshua Kimani, Keith R Fowke, Dawit Wolday, Charu Kaushic

**Affiliations:** Department of Biochemistry and Biomedical Sciences, Faculty of Health Sciences, McMaster University, Hamilton, ON, Canada; McMaster Immunology Research Centre, Faculty of Health Sciences, McMaster University, Hamilton, ON, Canada; Michael G. DeGroote Institute for Infectious, Diseases, McMaster University, Hamilton, ON, Canada; Cell Biology Program, Sloan Kettering, Institute, New York, New York, USA; Centre for Microbial Chemical Biology, Faculty of Health Sciences, McMaster University, Hamilton, ON, Canada; Department of Medical Microbiology and Infectious Diseases, University of Manitoba, Winnipeg, Manitoba, Canada; Department of Medical Microbiology, University of Nairobi, Nairobi, Kenya; Department of Medicine, Faculty of Health Sciences, McMaster University, Hamilton, ON, Canada

**Keywords:** Lipidomics, hormonal contraceptives, LC-MS, phospholipids, sphingolipids, HIV susceptibility, microbial lipids, pathway analysis

## Abstract

Heterosexual genital HIV transmission is a major driver of new infections, particularly in women, making them disproportionately vulnerable to HIV acquisition. Previous studies have associated injectable hormonal contraceptives (HC) with increasing susceptibility to HIV. Yet, the underlying molecular mechanism remains incompletely understood. Given the structural and signaling role of lipids in the female genital tract, cervicovaginal lipidomic profiling has the potential to reveal the mechanistic interplay among HC, lipidome, and HIV susceptibility in the female genital tract.

We conducted untargeted cervicovaginal lipidomics study in a cohort of high-risk, HIV-negative, Kenyan sex workers who were using injectable depot medroxyprogesterone acetate (DMPA), oral contraceptive pill (OCP), or no hormonal contraception (NH). Genital lipids were quantitatively analyzed using liquid chromatography-mass spectrometry (LC-MS) and bioinformatics platforms.

A total of 1045 lipid species were identified in the cervicovaginal lavage samples. Injectable DMPA significantly downregulated major structural and signaling membrane lipids, including phospholipids, ceramides, sphingomyelins, and glycosphingolipids (p<0.001, FDR<0.05), and markedly upregulated storage lipids (triglycerides) and sterols when compared with OCP or NH groups. Interestingly, microbial communities contributed significantly to the DMPA-driven genital lipidomic shifting, suggesting the intricate interplay among sex hormones, microbiome, and lipidome. In contrast, OCP, which contains estrogen, consistently lowered genital triglycerides and increased the fatty acid and diacylglycerol substrates compared with the NH group, implying that estrogen affects genital lipid metabolism by contrasting the lipogenesis effect of DMPA. Differential lipidomic pathway analysis further confirmed that DMPA suppressed the de novo phospholipid and sphingolipid biosynthesis, but enhanced salvaging of various lipids from reaction intermediates and from cell membrane lipid pool, shifting them towards TG accumulation.

Our findings of dysregulated depletion of several membrane and signaling phospholipids, ceramides, sphingomyelins, glycosphingolipids, along with upregulation of triglycerides in the DMPA users provides insights into the underlying molecular mechanism by which DMPA may increase in susceptibility to HIV infection by weakening the epithelial barrier and increasing chronic inflammation. These findings underscore the need to address cervicovaginal lipidomic dysregulation associated with HCs while designing interventions to improve women’s reproductive health.

## 1. INTRODUCTION

Heterosexual genital HIV transmission is a major driver of new infections, particularly in women, making them disproportionately vulnerable to HIV acquisition and associated co-morbidities ^1^. The female genital tract (FGT) is a highly integrated ecosystem where the microbes, metabolites, vaginal epithelium, local immunity, and sex hormones constantly interact to maintain its reproductive functions while balancing protection against pathogens ^2,3^. Disruption to any one of the components can destabilize the system and increase susceptibility to infections such as bacterial vaginosis (BV) and sexually transmitted infections (STIs) ^3–5^. Several studies, including fromour group, have shown the association of the injectable hormonal contraceptive (HC), depot medroxyprogesterone acetate (DMPA) with increased microbial diversity, hypo-estrogenism, thinning of the vaginal epithelial barrier, and inflammation, potentially increasing women’s susceptibility to HIV-1 ^5–8^. However, the underlying molecular mechanism for these off-target effects remains under-investigated.

Metabolomics, the comprehensive study of the metabolome, is the closest omics field to biological pathways, since metabolites reflect the most integrated and real-time snapshot of cellular processes, interactions in the local environment, and response to interventions ^9–12^. Specifically, lipid metabolites have emerged as potentially important in the FGT not only by indicating the state of host-microbial homeostasis ^13^ but also as markers predicting preterm birth ^14^, genital inflammation, and cervical cancer ^15^. Being a crucial component of cellular membranes, signaling pathways, immune mediators, and energy storage, lipids form the structural foundation of the FGT tissues, maintain cellular integrity, modulate pathogen entry, regulate immune and inflammatory responses, and fuel the bioenergetics of host cells and the microbial community ^16–19^. While microbe-derived lipids evidently impact host lipidome in the gut ^20^, little is known about the factors that affect the genital lipidome and their role in the FGT.

Hormonal contraceptives (HC), including the injectable DMPA, are among the most commonly used birth control methods in sub-Saharan Africa and across the globe ^21,22^. However, to date, there is no data showing how hormonal contraceptives shape the lipid metabolism in the genital tract and if that is associated with altered susceptibility to infection in women. To address this gap, we performed untargeted cervicovaginal lipidomics study among high-risk, HIV-negative women sex workers who were using different hormonal contraceptives. With the aim of identifying specific lipid signatures associated with use of injectable DMPA or oral hormonal contraceptive pills (OCPs), we quantitatively profiled the lipidomic landscape of the FGT using liquid chromatography-mass spectrometry (LC-MS)-based lipidomics and bioinformatics analysis. Beyond identifying individual lipid signatures, we further mapped the mechanistic links among lipid metabolic pathways as potential molecular mechanisms. Understanding how HCs impact genital lipid metabolism will contribute to addressing off-target effects while designing interventions to improve women’s reproductive health.

## 2. METHODS

### 2.1. Study subjects and cervicovaginal lavage samples

The participants in this study were all adult women with heterosexual gender identity. They were engaged in sex work and recruited from the Sex Workers Outreach Programme (SWOP) and Pumwani Sex Worker cohort in Nairobi, Kenya. Screening and enrollment procedures were conducted in accordance with all ethical guidelines up on approval by the Research Ethics Boards of McMaster University (HiREB 0332-T,), University of Manitoba (B2015:033) and Nairobi/Kenyatta National Hospital (KNH-ERC P132/03/2015). Participants were enrolled based on the inclusion and exclusion criteria previously described ^5^. Briefly, women were included if >18 years old, pre-menopausal, non-pregnant, not <1 year of breastfeeding, willing to undergo pelvic exams, intact uterus and cervix, in good health and negative for STIs (gonorrhea, chlamydia, Trichomonas vaginalis, syphilis, HIV), negative for yeast infection, Nugent score <7 at screening, could abstain from douching for 24 h and sexual activities for 12 h before sample collection, use condoms for 36 h prior to abstinence, and had not been in sex work >5 years. From a total of 370 sex workers screened, 45 women who completely fulfilled the inclusion criteria could be recruited for this study. From these, 20 sex workers were taking injectable depot medroxyprogesterone acetate (DMPA) for >6 months and screening was within 3-4 weeks of the most recent injection, 10 sex workers were on oral contraceptives pills (OCPs) containing estrogen and progesterone for > 6 months and screening was within 5-10 days of beginning a new pack, 15 sex workers were not taking any hormonal contraception (NH) for >6 months and screening was within 5-10 days of beginning a menstrual cycle (proliferative phase). After carrying out all relevant gynaecological exams, cervicovaginal lavage (CVL) samples were collected from the posterior vaginal fornix of the selected participants and then placed in a sterile tube on ice and sent to the laboratory, where it was centrifuged at low speed (120 g) to remove cellular debris. CVL supernatants were frozen and stored at −80°C until shipped to Winnipeg, Manitoba, Canada, after which carefully shipped on dry ice to Hamilton, Ontario, Canada, and again stored at −80°C until lipidomic analysis was carried out.

### 2.2. Lipid extraction from cervicovaginal lavage

Frozen CVL samples, including 20 samples from DMPA, 10 samples from NH, and 15 samples from NH groups, were first thawed at room temperature. Then, 20 µL (optimized through pilot experiments) of each sample was separately transferred into labeled 1.5-mL polypropylene Eppendorf tubes, and 10 µL of deuterated lipid internal standards (IS) mixture (Avanti SPLASH LipidoMIX™ Internal Standard, prepared by diluting the stock 3x with methanol) was added to each tube. The multiple phospholipids, glycerolipids, sphingolipids, and cholesterol lipid subclasses from the IS mixture are used for normalization of their respective lipid species identified from the samples. Similarly, 10 µL of 100 µg/mL deuterated Docosahexaenoic Acid; DHA-d5 (Cayman chemical) was also added to be used for normalization of free fatty acid species. Next, proteins were precipitated upon addition of cold methanol (1:4 sample: methanol, v/v) with vortexing and centrifuging (15,000 rpm, 30 min, 4 °C), followed by transferring the supernatant to newly labeled Eppendorf tubes. Each mixed solution was then extracted by a modified MTBE lipid extraction method, as previously described ^23^. Briefly, lipids were extracted by adding 260 µL methyl *tert*-butyl ether (MTBE) (1:13 sample: MTBE, v/v), vortexing, and incubating for 1 hour at 4 °C with gentle shaking. Phase separation was induced by adding 70 µL of MS-grade water (1:3.5 sample: water, v/v), vortexing, and incubating ((15,000 rpm, 10 min, 4 °C). The upper lipid-containing organic phase was carefully transferred to a new labelled tube, and the lower layer was re-extracted with MTBE/methanol/water (10:3:2.5, v/v/v) using a 1:10 sample: solvent ratio and following similar steps described above. The combined lipid layer was concentrated using a Turbovap^®^ nitrogen evaporator (Biotage, Uppsala, Sweden), and then dried lipid extracts were reconstituted with 60 µL of MeOH/isopropanol (1:1, v/v). Thereafter, a pooled QC sample (prepared by mixing 5 µL from all CVL samples) and the remaining 55 µL of each sample were finally transferred to labelled HPLC vials with glass inserts for LC-MS analysis. To assess variability of the extraction method through internal standard recovery, three CVL samples from the NH group were extracted in duplicate, with IS added before extraction in one replicate and after extraction in the other.

### 2.3. Lipidomic analysis using LC-MS method

The separation and data acquisition of extract lipids was performed using reversed-phase ultra-performance liquid chromatography coupled to quadrupole time-of-flight mass spectrometry (UPLC/Q-TOF MS). The liquid chromatography (LC) system consisted of an Agilent 1290 series UPLC (Agilent Technologies, Santa Clara, CA, USA) equipped with Agilent C18 (2.1 × 100 mm, 1.8 μm) column for the separation of lipid mixtures. Autosampler and column temperatures were maintained at 10°C and 60 °C, respectively. To maximize the number of detected lipid species, MS analysis was performed in full-scan mode in both positive and negative ionization modes over an m/z range of 100-1,700 amu. The mobile phases consisted of mobile phase A and mobile phase B. Mobile phase A was composed of 60:40 (v/v) acetonitrile/water with 10 mM ammonium formate and 0.1 % formic acid for positive ion mode and 60:40 (v/v) acetonitrile/water with 10 mM ammonium acetate for negative ion mode. Mobile phase B consisted of 90:10 (v/v) isopropanol/acetonitrile with 10 mM ammonium formate and 0.1% formic acid for positive mode and 90:10 (v/v) isopropanol/acetonitrile with 10 mM ammonium acetate for negative mode. The LC gradient used was 15% to 99% solvent B over 12 min, with a return to 15% B and a final isocratic hold at 15% B prior to the next injection. The flow rate was 0.6 mL/min, with the injection volume of 2μL and 5μL for positive and negative ion modes, respectively. LC-MS grade isopropanol was used as the needle wash solvent to minimize the intensity of the background signal. The MS detection of separated lipids was performed using a Q-TOF/MS (Agilent Technologies, Santa Clara, CA, USA) equipped with an electrospray ionization (ESI) source. Instrument parameters were identical for both positive and negative ion modes as follows. Drying gas temperature 200°C, drying gas flow 13L/min, nebulizer pressure 35 psi, sheath gas temperature 350 °C, sheath gas flow 11L/min, capillary voltage ±3.5 kV, nozzle voltage 1 kV, and acquisition rate 2 spectra/s. Prior to sample analysis, external mass calibration was performed using a standard calibration mixture. Reference masses of m/z 922.009798 and 121.050873 for positive mode, as well as 112.9855 and 1033.9881 for negative mode were used and continuously monitored throughout the analytical run as lock masses to ensure and maintain mass accuracy.

A pooled quality control (QC) sample, prepared by blending aliquots of all individual sample extracts, was injected every five samples to monitor instrument performance across the analysis. Solvent blank, composed of MeOH/isopropanol (1:1, v/v), was injected at the beginning of the sequence, before and after QC injections, and among samples of different study groups to avoid carryover and cross contamination. Data acquisition was performed using Agilent MassHunter Workstation software (Agilent Technologies, Santa Clara, CA, USA).

### 2.4. Lipidomic data processing and curation

Detection profile and recovery of deuterated lipid internal standards were inspected by Agilent MassHunter Qualitative Analysis software (Agilent Technologies, Santa Clara, CA, USA) using their respective total and extracted ion chromatograms (TIC/EIC), retention times (RT), and mass spectra. The recovery of each internal standard was assumed to be representative of the recovery for related lipid species in that class or subclass. Ion adducts included +H, +NH_4_^+^, +Na^+^ in positive ion mode and –H, +HCOO^−^ in negative ion mode. Agilent MassHunter Profinder 10.0 software (Agilent Technologies, Santa Clara, CA, USA) was used for batch feature extraction and chromatographic alignment from Q-TOF/MS acquired data, playing a critical step in the peak finding and data reduction workflow. The feature files generated were then exported as Profinder Archive (PFA) files to Agilent Mass Profiler Professional (MPP). MPP enabled group wise sorting and visualization of differentially expressed peak intensities of compound features. One key feature of the MPP is being integrated with Metabolite and Chemical Entity (METLIN) database (version 2025) and Human Metabolome Database (HMDB) that enabled putative annotation of the compound features to a wide range of lipid metabolites using their characteristic m/z, RT, and isotope patterns. Identified lipid data was finally normalized using corresponding lipid class internal standards and exported as CSV file for further statistical analysis. Lipids that showed missing values in ≥ 30% of the study subjects, had >25 % CV in the pooled QC replicates, or lacked an appropriate IS for lipid class-based normalization were considered imprecise and subsequently excluded.

### 2.5. Statistical analysis

Lipid relative abundances were log₂-transformed and z-score normalized (mean = 0, SD = 1) to enable direct comparison of effect sizes across lipid species, such that regression coefficients represent the change in outcome per one standard deviation increase in lipid abundance. Cross-sectional associations between the vaginal lipidome and contraceptive use were evaluated using multinomial logistic regression with three outcomes groups: DMPA users, OCP users, and NH users, after adjusting for age and alcohol use. Multiple comparisons were addressed using complementary correction strategies: Bonferroni correction for discovery phase filtering (p < 1 × 10⁻⁵; α = 0.05/1045 lipids) and Benjamini–Hochberg false discovery rate control for biological interpretation (FDR < 0.05). Volcano plots visualized effect sizes (log₂ odds ratios) against statistical significance (-log₁₀ FDR-adjusted p-values). Spearman rank correlation was used to examine relationships among significant lipids (FDR < 0.05), with results visualized as hierarchical clustering heatmaps (Euclidean distance, Ward’s D2 linkage). Principal component analysis (PCA) was performed on standardized abundances to assess global lipidomic patterns by contraceptive group. Lipid species were classified using the LIPID MAPS® database ^24^ and used as the reference background for enrichment analyses to avoid pre-filtering bias. Lipid class enrichment was assessed using the Lipid Ontology (LION) platform ^25^ and biological pathway analysis was performed using BioPAN, with LIPID MAPS-curated metabolic pathways. Statistical analyses and visualization were performed using R v4.3, MetaboAnalyst v6.0, or GraphPad prism v10.6.1.

## 3. RESULTS

To uncover the effect of DMPA and OCP on lipid metabolism in the genital tract and examine their potential molecular mechanisms that influence susceptibility to HIV infection, we performed untargeted lipidomics study using 45 CVL samples taken from Kenyan female sex workers belonging to 20 DMPA, 10 OCP, and 15 NH user groups. The samples were from a previous study where participants were screened using thorough selection criteria involving questionnaire, lab tests, and clinical investigations. While detailed demographic information has been reported in the previous parent study ^5^, there was no significant difference in age or any other demographic information among the three study groups (P > 0.05) other than the use of DMPA, OC or NH.

### 3.1. Genital lipidome landscape

Overall, a large-scale lipidomics workflow was optimized (Figure 1) which enabled the detection of 3635 features in positive ion mode and 1511 in negative ion mode experiments. Further lipid annotation using the METLIN and HMDB platforms resulted in the identification of 1560 lipids (42.9%) in positive ion mode and 902 lipids (59.7%) in negative ion mode. This incomplete identification, with a large proportion of detected featured being labeled as unknown shows that lipidomics and associated bioinformatics databases are still in early stages and lagging behind other omics fields. We further narrowed the lipid list to a total of 1045 based on data normalization, QC (% CV), and missing value filtering criteria.

**Fig. 1:**
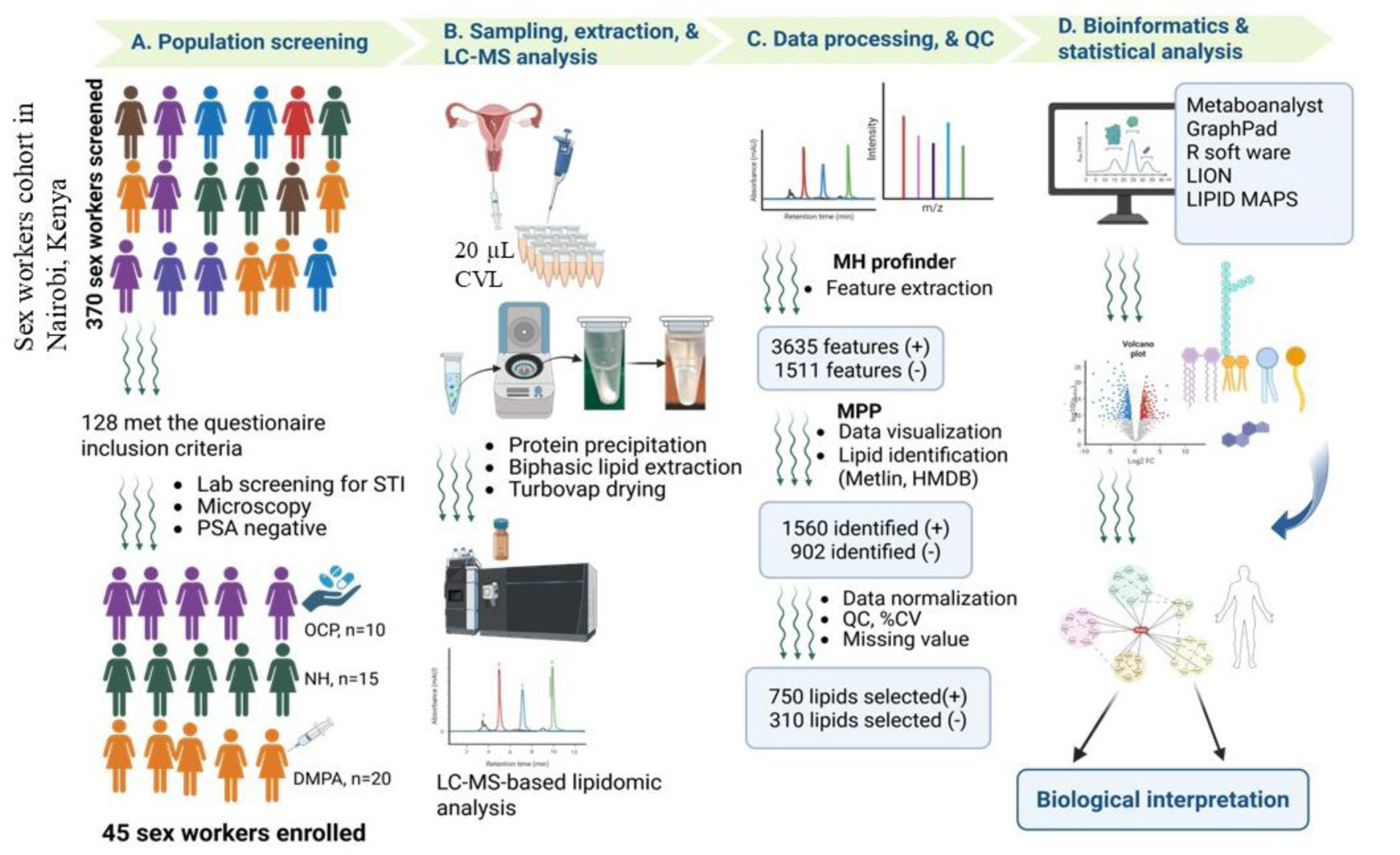
Study design and LC-MS-based workflow for cervicovaginal lipidomic analysis.

The identified 1045 CVL lipid species belong to fifteen main lipid subclasses, with triglycerides (TG) (30%), phosphatidylserine () (9.3%), free fatty acid (FFA) (9.0%), phosphatidylcholine (PC) (8.5%), and phosphatidylethanolamine (PE) (6.2%), accounting for the top five lipid components (Fig. 2A). This lipid distribution, describing the relative contribution of each lipid subclass to the total lipidome, highlights the predominance of lipid storage, energy source, and membrane-associated lipid species within the genital lipidome landscape. The normalized lipid abundance dataset of the identified lipids was used for further quantitative statistical analysis. To examine the differences in the whole lipidome among the three study groups, we first performed a principal component analysis (PCA) using the entire lipidome dataset. The PCA plots illustrate a clear distinction between the study groups based on the total lipidome (Fig. 2B-D). Specifically, injectable and oral hormonal contraceptive users show distinct lipidomic profiles, while the NH group occupied an intermediate position between DMPA and OCP clusters (Fig. 2E), suggesting differential lipidomic signatures associated with use of specific contraceptives.

**Fig. 2:**
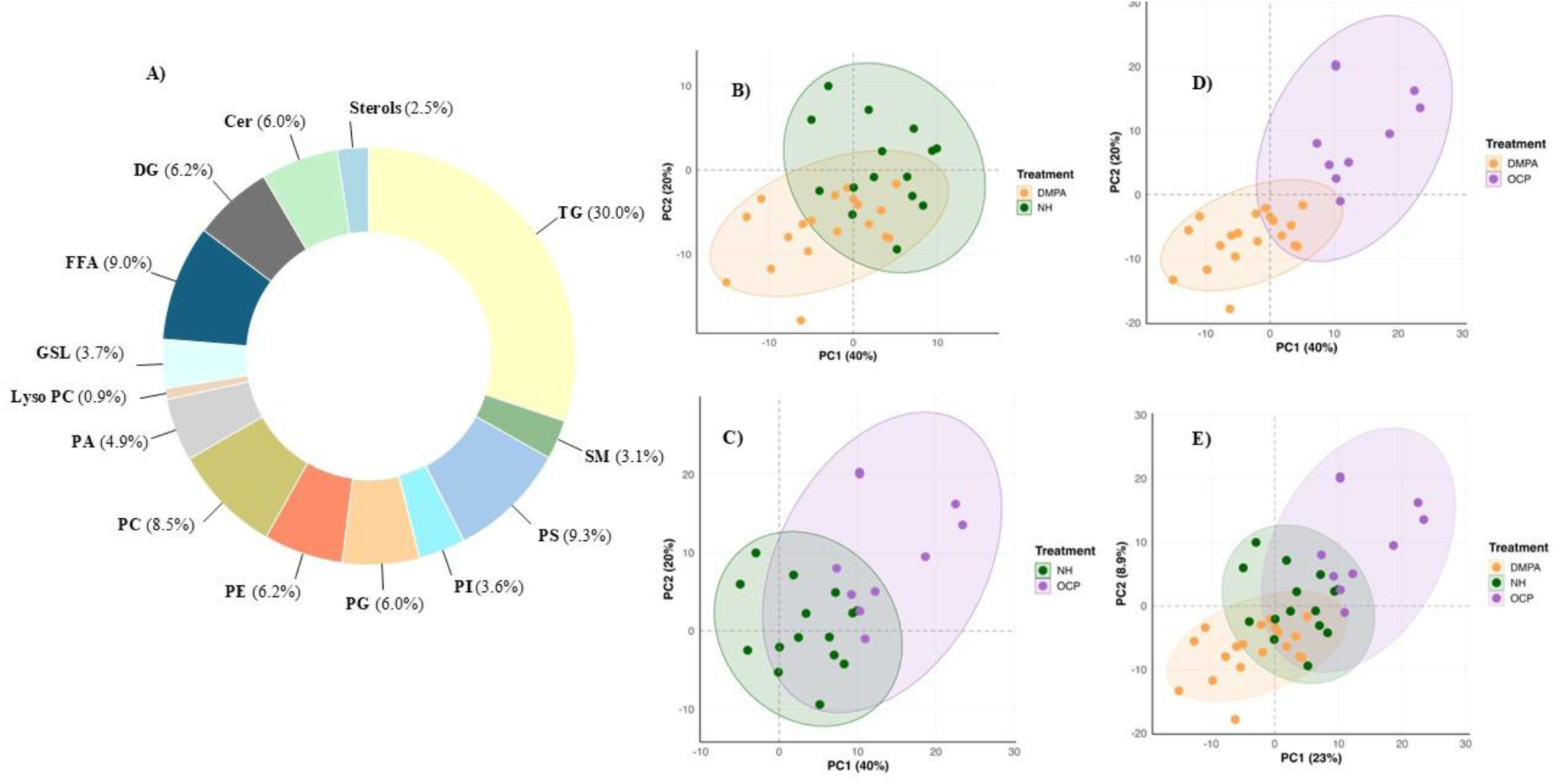
Cervicovaginal lavage lipidome profile across study groups. **A**) Donut chart showing overall lipidome composition among major lipid sub-classes. Principal component analysis (PCA) score plot of whole lipidome between: **B**) DMPA and NH; **C**) NH and OCP; **D**) DMPA and OCP groups; **E**) DMPA, OCP, and NH groups. Each point represents an individual sample, and ellipses denote the multivariate dispersion of each group. DMPA: Depot Medroxyprogesterone acetate; NH: no hormone; OCP: oral contraceptive pill

### 3.2. DMPA use downregulated structural and signaling lipids while elevating storage lipids compared to women who were on OCP or did not use HC

To further explore the lipid species driving the noted lipidomic changes, we next examined lipids that were significantly altered among the study groups using ANOVA-based analysis. Fisher’s LSD based ANOVA test of the CVL lipidome revealed 333 lipid species showing significant differences (false discovery rate, q ≤ 0.05) among the study groups. Lipids showing distinct differences were visualized using bar graphs, heatmaps, volcano plots, and were classified according to their major lipid subclasses. The total lipidome abundance of major membrane phospholipids illustrate significant (PA, PC, PE, lyso-PC) and non-significant (PS, PI, PG, and CL) depletion in the DMPA users compared to the NH or OCP users (Fig. 3A, B). Among these lipidome subclasses, 36 phosphatidylcholine (Fig. 3C), 13 phosphatidylethanolamine (Fig. S1A), and 18 other phospholipid molecular species (Fig. 3D) demonstrated significant downregulation in the DMPA use group.

**Fig. 3:**
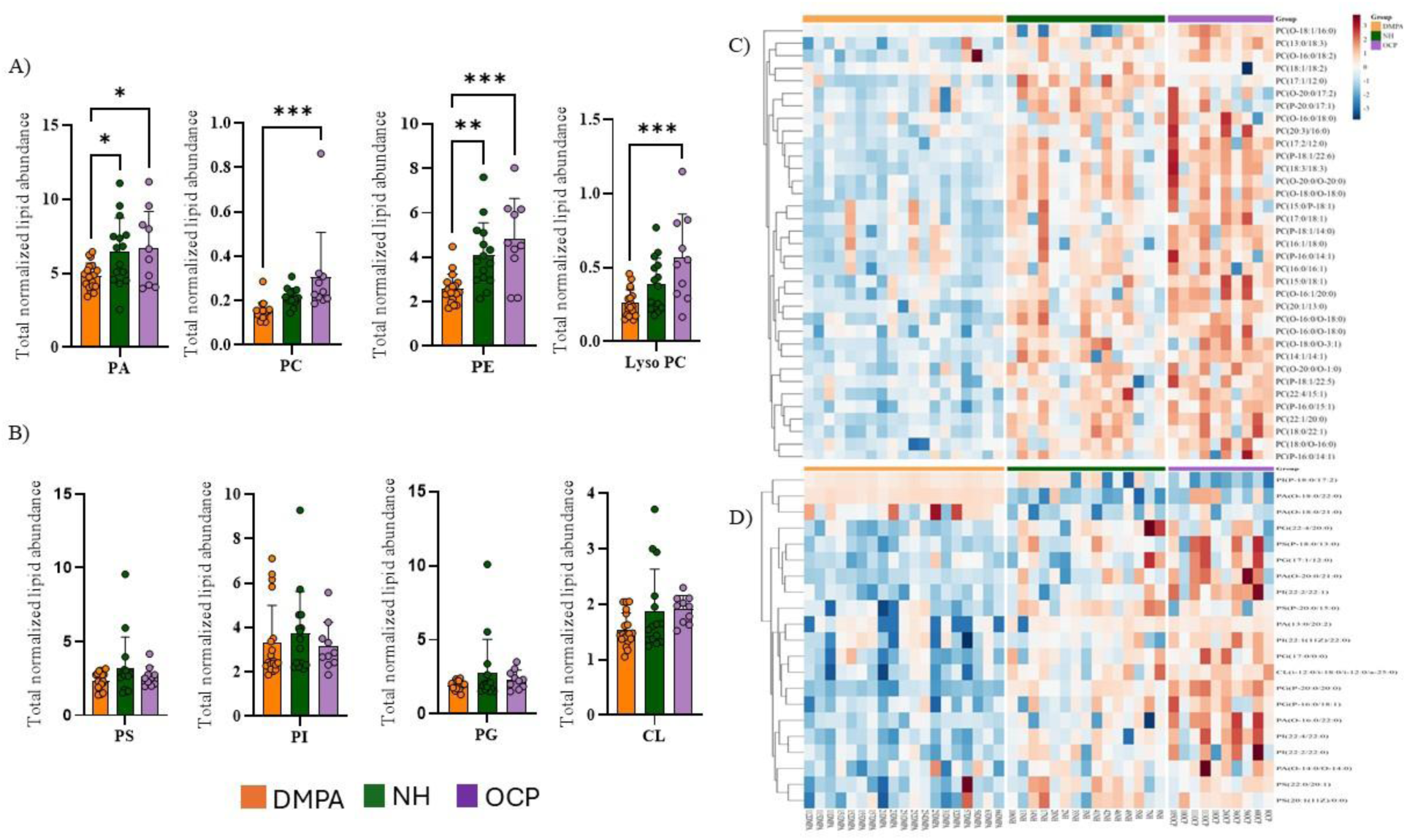
Differentially altered genital phospholipids among DMPA, NH, and OCP taking groups. **A**) Significantly changed total lipid abundance of phospholipid sub-classes (PA: phosphatidic acid, PC: phosphatidylcholine, PE: phosphatidylethanolamine). **B**) Non-significantly changed total lipid abundance of other phospholipid sub-classes (PS: phosphatidylserine, PI: phosphatidylinositol, PG: phosphatidylglycerol, CL: cardiolipin). **C**) Heatmap showing individual phosphatidylcholine (PC) species markedly downregulation (*q* ≤ 0.05) in the DMPA taking group. **D)** Heatmap showing individual other phospholipid species (PA, PS, PG, PI, CL) markedly downregulation (*q* ≤ 0.05) in the DMPA taking group. Rows represent individual lipid species and columns correspond to study groups: depot medroxyprogesterone acetate (DMPA), no hormone (NH), and oral contraceptive pill users (OCP). Heatmap color intensity represents scaled lipid abundance (z-score), where red indicates relatively higher abundance and blue indicates lower abundance across groups. Asterisk (*) shows level of significance using adjusted *p*-value or false discovery rate (*q*) in which *** means 0.0001 < *q* ≤ 0.001, ** means 0.001 < *q* ≤ 0.01, * means = 0.01<q ≤0.05.

Similarly, the prominent membrane sphingolipids including ceramides, sphingomyelin, and glycosphingolipids showed a significant reduction in total lipid abundance across these subclasses (Fig. 4A) and in specific glycosphingolipid (Fig. 4B), sphingomyelin (Fig. 4C), and ceramide (Fig. 4AD) species in DMPA users compared with OCP or NH user groups. These sphingolipids, along with the phospholipids and are known to represent structural and signaling lipids of biological membranes in both host and microbial species.

**Fig. 4:**
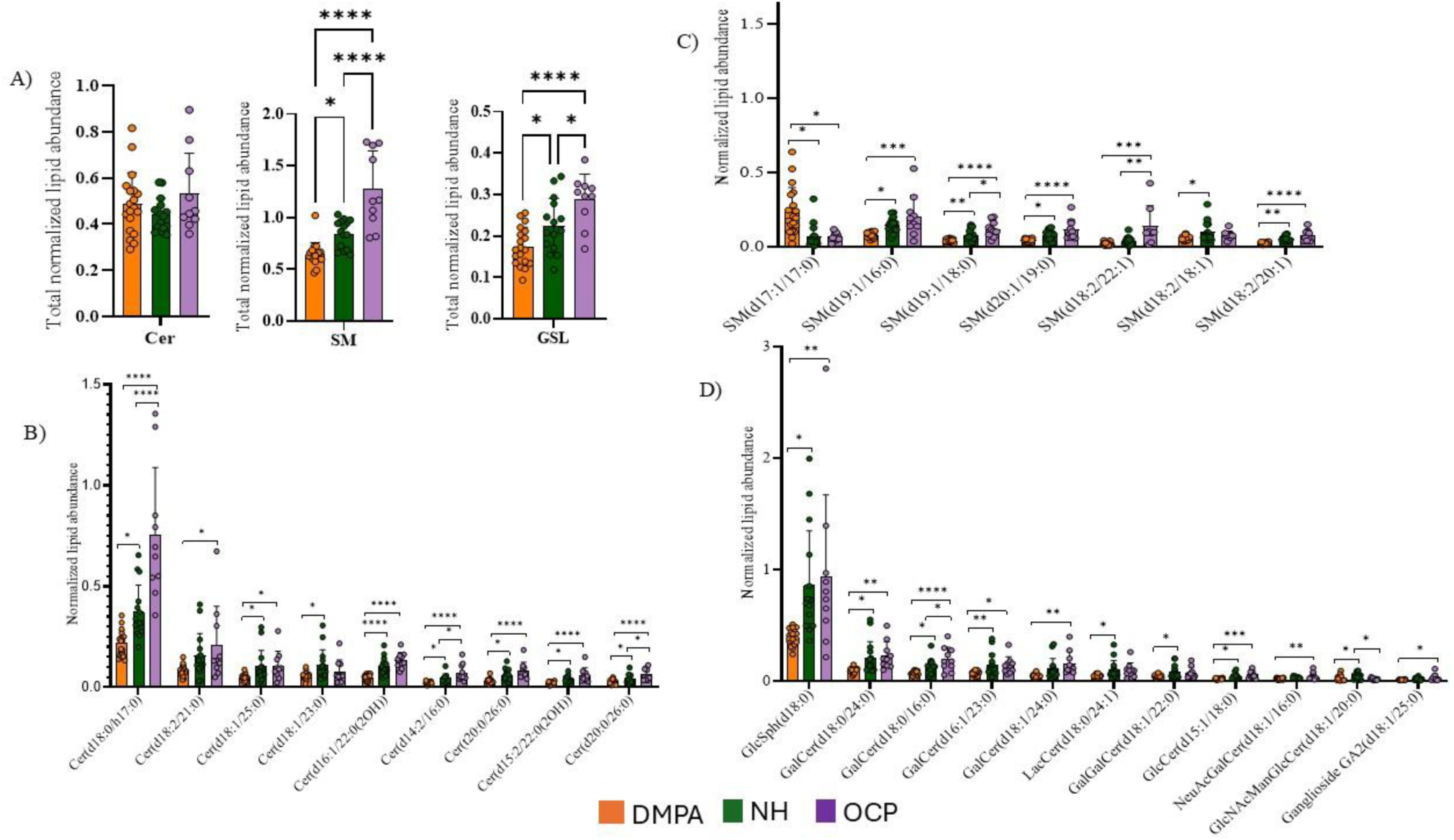
Differentially altered genital sphingolipids among DMPA, NH, and OCP taking groups. **A**) Total lipidome abundance of three sphingolipid sub classes: Ceramide (Cer), Sphingomyelin (SM), and Glycosphingolipid (GSL). **B**) Significantly altered specific ceramide species. **C**) Significantly altered specific sphingomyelin species. **D**) Significantly altered specific glycosphingolipid species. Data are expressed as mean ± SD. **** means *q* ≤ 0.0001, *** means 0.0001 < *q* ≤ 0.001, ** means 0.001 < *q* ≤ 0.01, * means 0.01< *q* ≤ 0.05.

On the other hand, differential shifts in neutral and storage lipids occurred in the opposite direction, such that the CVLs from women using injectable DMPA contraceptive showed consistent upregulation of the triglycerides (Fig. 5A, C, D) and sterol lipids (Fig. 5B and Fig S1B) when compared with the OCP and NH groups.

**Figure 5:**
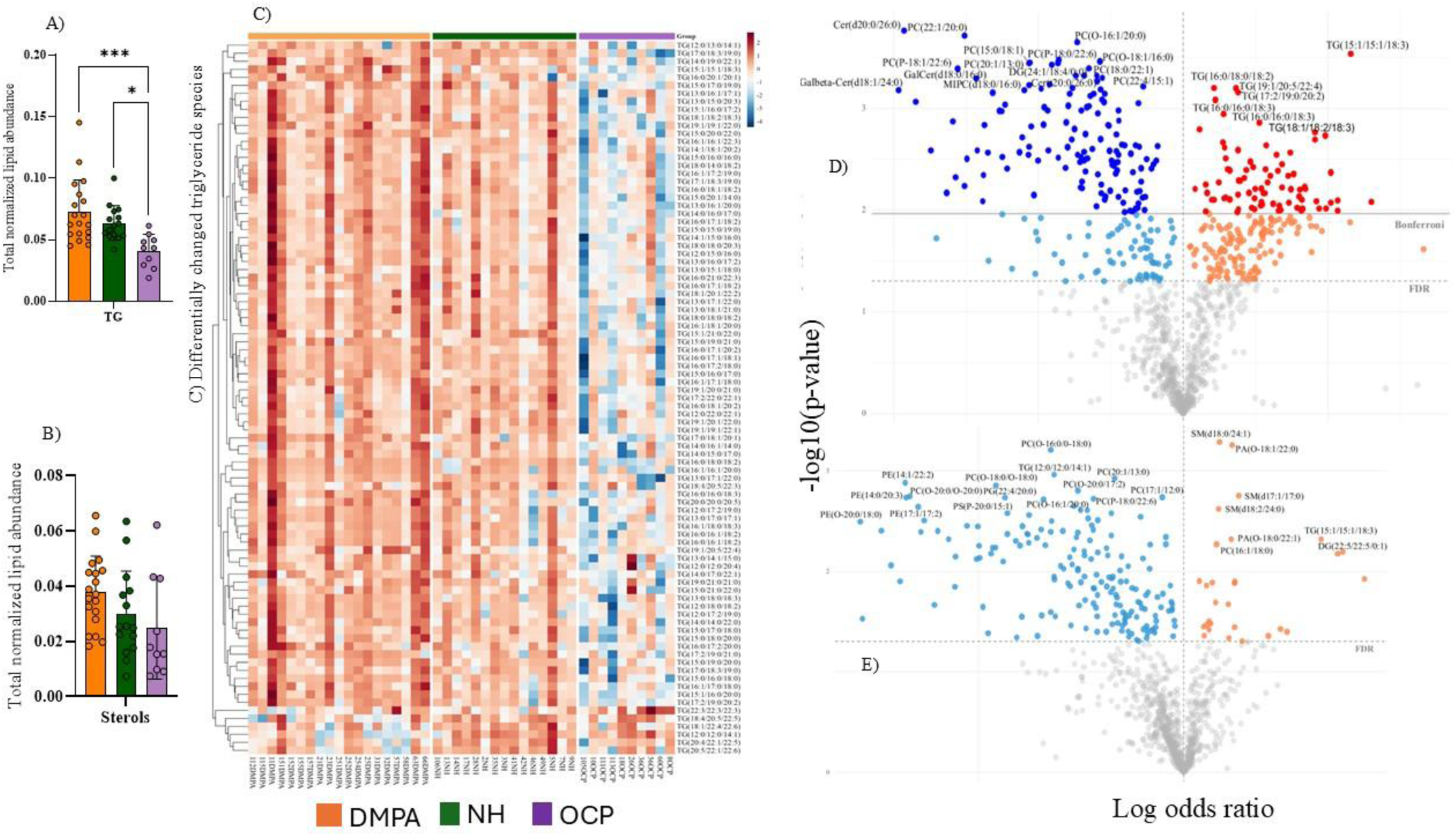
Differentially dysregulated storage lipids and sterols among the study groups. **A**) Differential abundance of triglycerides (TG) subclass. **B**) Differential abundance of sterol subclass. **C**) Heatmap illustrating significantly (*q* ≤ 0.05) upregulated specific TG species in the DMPA group. **D**) Volcano plot of genital lipidome distribution illustrating DMPA associated upregulation of TG species and downregulation of most other lipid species when compared with the OCP group. **E)** Volcano plot illustrating genital lipidome changes associated with DMPA compared with NH group. Pairwise comparisons are based on logistic regression models using log-transformed odds ratios. Each point in the volcano plot represents an individual lipid species plotted according to effect size and statistical significance, with intense blue: highly downregulated (*q* ≤ 0.05) in DMPA, blue: downregulated (*p* ≤ 0.05) in DMPA, red: highly upregulated (*q* ≤ 0.05) in DMPA, orange: upregulated (*p* ≤ 0.05) in DMPA, grey: not significant. *** means 0.0001 < *q* ≤ 0.001, * means 0.01< *q* ≤ 0.05.

### 3.3. OCP use depleted triglyceride but enhanced its metabolic precursors

CVLs from women who were on OCP, which is a combination of estrogen and progestin formulations, showed marked depletion of the storage lipid TG (Fig. 5C and Fig. 6D) and upregulation of its metabolic precursors, FFA and diacylglycerol (DG) (Fig. 6A-C and Fig. S2A), reshaping the lipidomic environment in the genital tract very differently and in contrast to DMPA. Similarly, most genital sterol lipids were seen to have lower abundance in the OCP group compared with the DMPA and NH groups (Fig. 5B and Fig S1B).

**Figure 6:**
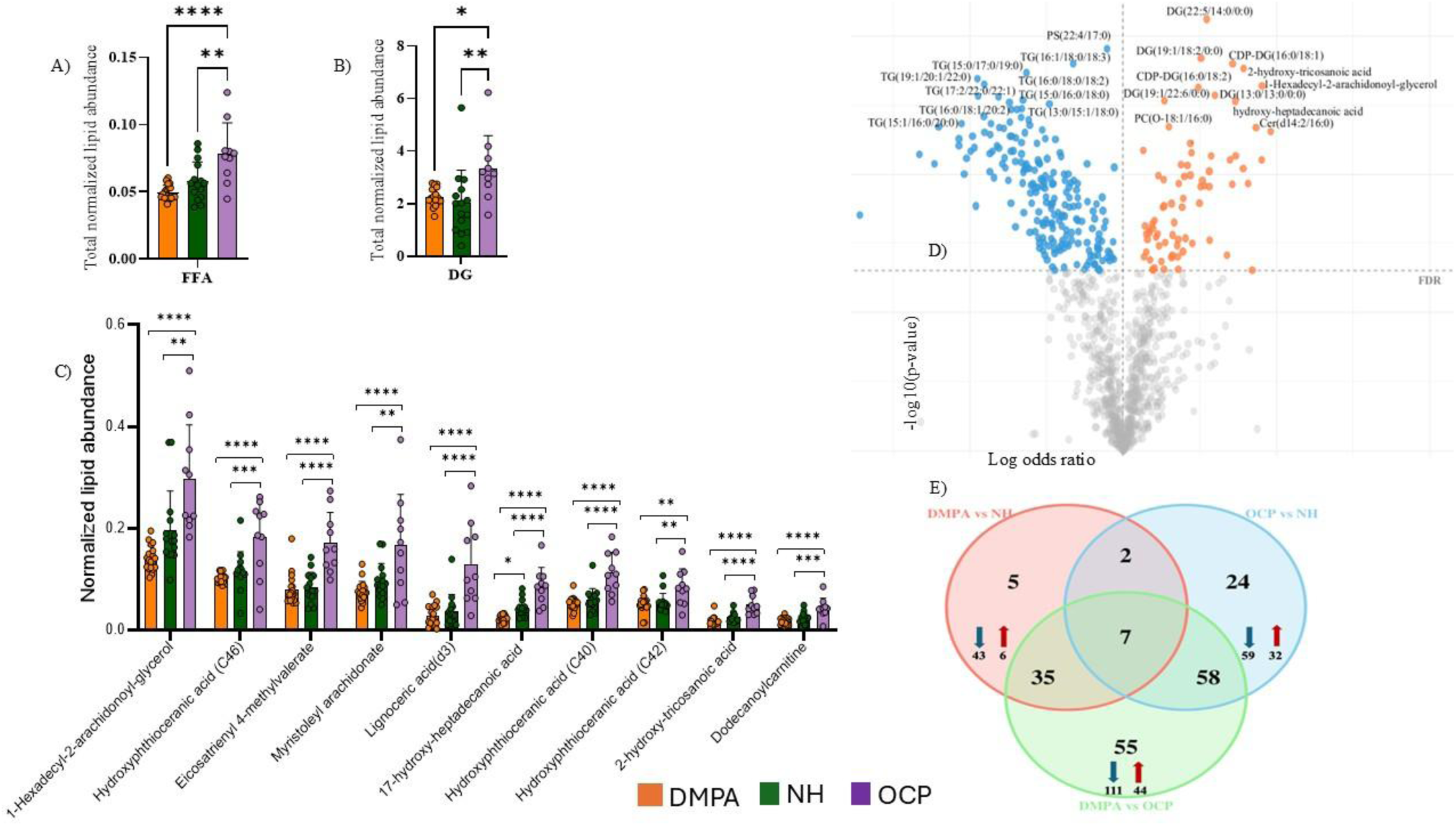
Differential lipidomic shifting among the study groups. OCP driven significant upregulation of total FFA subclass (**A**), total DG subclass (**B**), specific FFA species (**C**). **D**) Volcano plot illustrating OCP associated downregulation of TGs and upregulation of FFAs and DG when compared with the NH group. **E**) Venn diagram showing unique and shared lipid signatures with their direction of shifting among pair wise comparisons. Each point in the volcano plot represents an individual lipid species plotted according to effect size and statistical significance, with blue: downregulated (*p* ≤ 0.05) in OCP, orange: upregulated (*p* ≤ 0.05) in OCP, grey: not significant. **** means *q* ≤ 0.0001, *** means 0.0001 < *q* ≤ 0.001, ** means 0.001 < *q* ≤ 0.01, * means 0.01< *q* ≤ 0.05.

We further explored the unique and shared lipid signatures among the study groups through pair wise comparisons. This was based on multinomial logistic regression analysis (log odd ratios) using Bonferroni correction (p < 0.05/number of detected lipids) as a selection criterion. As summarized in Fig. 6E, the pair wise comparison between DMPA and OCP users exhibited the largest number of changes in 155 lipid species, within which 55 lipids were uniquely dysregulated, while 58 and 35 dysregulated lipid signatures were shared with the OCP vs NH and DMPA vs NH comparisons, respectively. As such, 7 other dysregulated lipids comprising of Cer(d18:0/h17:0), MIPC(t18:0/16:1), PC(20:1/13:0), PC(22:1/20:0), PC(O-16:1/20:0), PC(P-18:1/22:6), and TG(15:1/15:1/18:3) were common to all pair wise comparisons.

To investigate how strongly or weakly each lipid is co-regulated with any other lipid, we performed pairwise spearman correlation analysis and found marked patterns of inter-lipid co-regulation. Notably, top 90 correlated lipids among the whole lipidome data consistently demonstrated negative correlation of triglycerides with most phospholipids, sphingolipids, and free fatty acids (Fig. S2B).

### 3.4. Microbially produced lipids accounted for the differential lipidomic shifting seen among women using DMPA or OCP

Since the differential changes observed in the cervicovaginal lipidome could be driven by both host and microbial lipid metabolism, we further wanted to estimate the microbe specific changes in lipid metabolism. To this end, we extracted and analysed the odd carbon lipid species, given that odd carbon chain fatty acid containing lipids are predominantly produced by microbial communities ^20,26^. As observed in the whole lipidome, the microbial membrane phospholipids and sphingolipids showed significant downregulation in the injectable DMPA user group when compared to the NH or OCP user groups (Fig. 7A, B). Consistent with the whole lipidome, microbe specific TGs also showed marked upregulation associated with DMPA contraception (Fig. 7C). Specifically, six microbially produced lipid sub-classes namely PC, TG, Cer, SM, DG, and glycosphingolipids (Fig. 7D) constituted the top thirty lipids that contributed to the lipidomic variation among the three study groups.

**Fig. 7:**
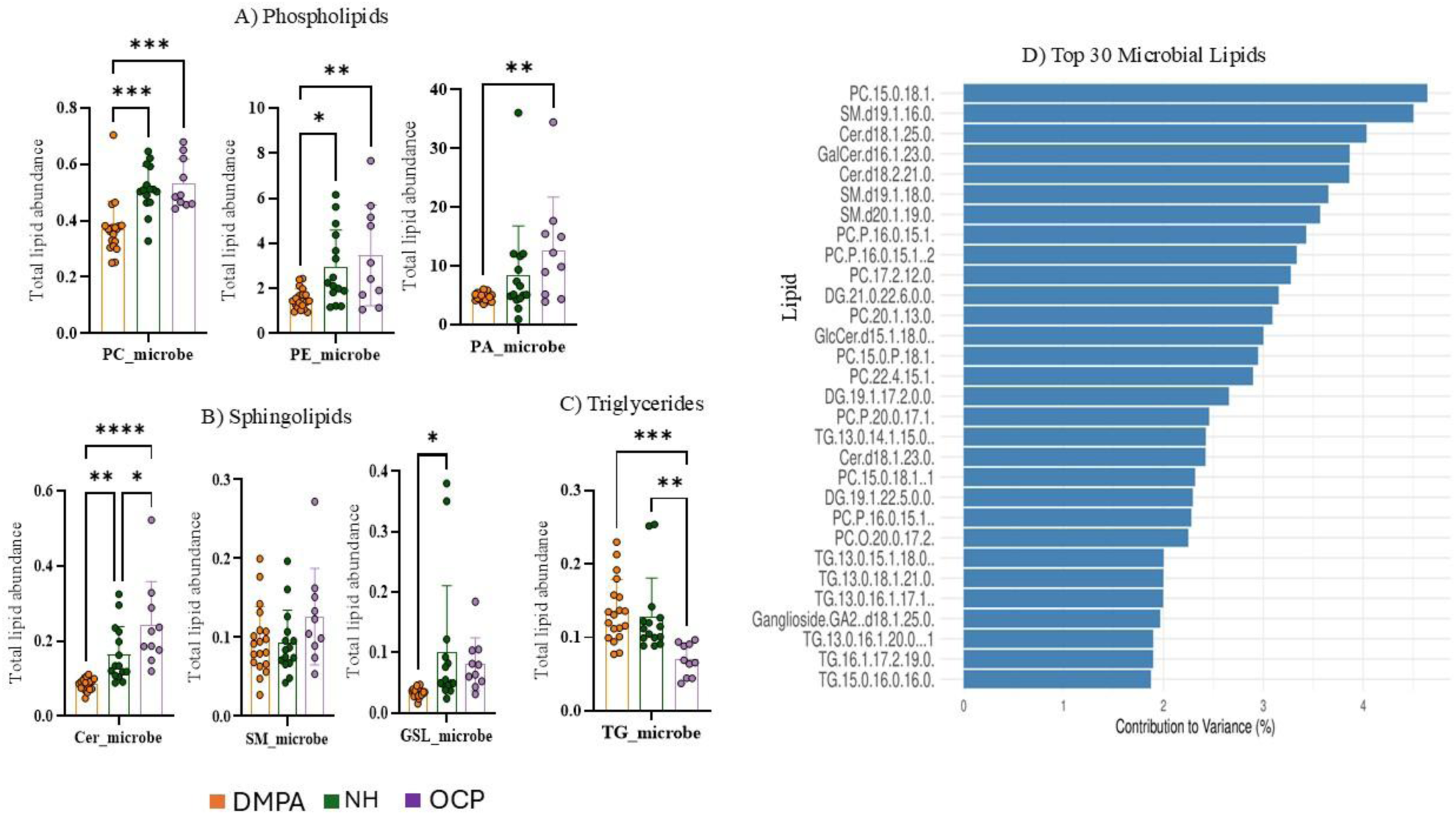
Microbially produced lipids exhibited hormonal contraceptive specific lipidomic shifting. Total microbial lipidome abundance of phospholipid (**A**), sphingolipid (**B**), and triglyceride (**C**) subclasses among the study groups. **D**) Top thirty microbial lipid species that caused the inter-group variation among DMPA, NH, and OCP taking groups.

### 3.5. Lipid enrichment and pathway analysis

We performed Lipid Ontology (LION) enrichment analysis using ranking mode to map the dysregulated lipids and provide further insight into their cellular location, function, chemical, biophysical, and biological properties. This approach identified over-represented lipid features by ranking among the significantly altered lipid species. The comparison among the different HC groups, revealed a strong enrichment of lipids belonging to storage and lipid droplets, neutral and charged head groups, membrane components, signaling groups, short and long chain fatty acids, sphingolipids, and endoplasmic reticulum at the top of the ranking mode (Fig. S3A-D). To examine the metabolic pathways affected by the DMPA driven lipidomic dysregulation, we used BioPAN integrated with LIPID MAPS tool. BioPAN uses a Z-score algorithm to predict whether specific metabolic reactions are activated (positive Z-score) or suppressed (negative Z-score). Pathway analysis (Fig. 8 and Table 1) clearly illustrated changes in lipid metabolism, with specific enhancement (highlighted in orange color and positive Z-score) of pathways leading to TG, mitochondrial cardiolipin, and membrane lipid salvaging. On the contrary, DMPA imparted significant suppression (highlighted in blue color and negative Z-score) of lipid pathways producing phospholipids and sphingolipids which are the major structural and signaling components of biological membranes.

**Fig. 8:**
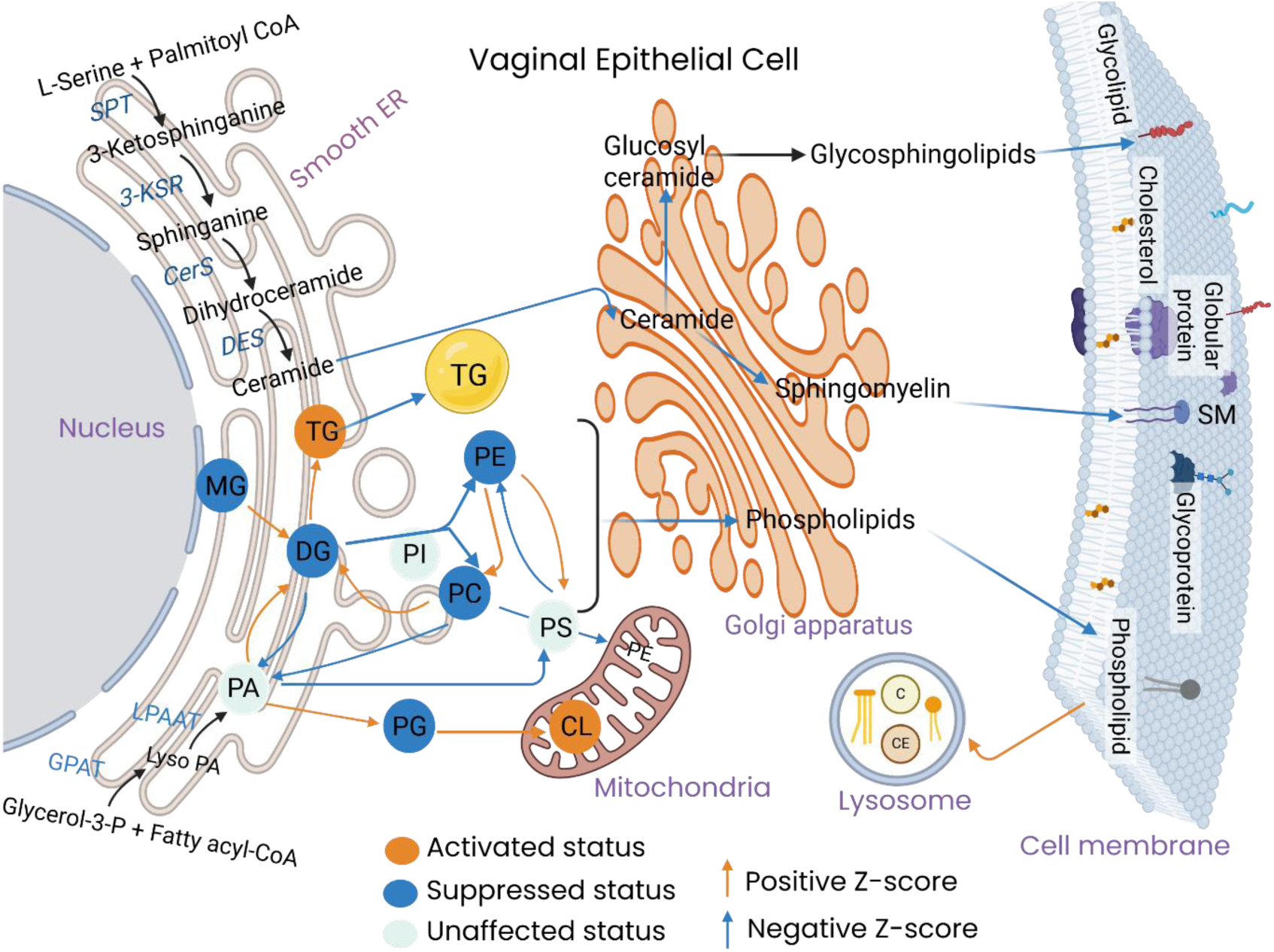
BioPAN based metabolic pathway changes associated with DMPA exposure in vaginal epithelial cell. This network illustrates the predicted metabolic shifts with nodes (bubbles) showing suppressed or activated (statistically significant change) lipid sub-classes and arrows depicting metabolic pathways. MG: monoacylglycerol.

**Table 1:**
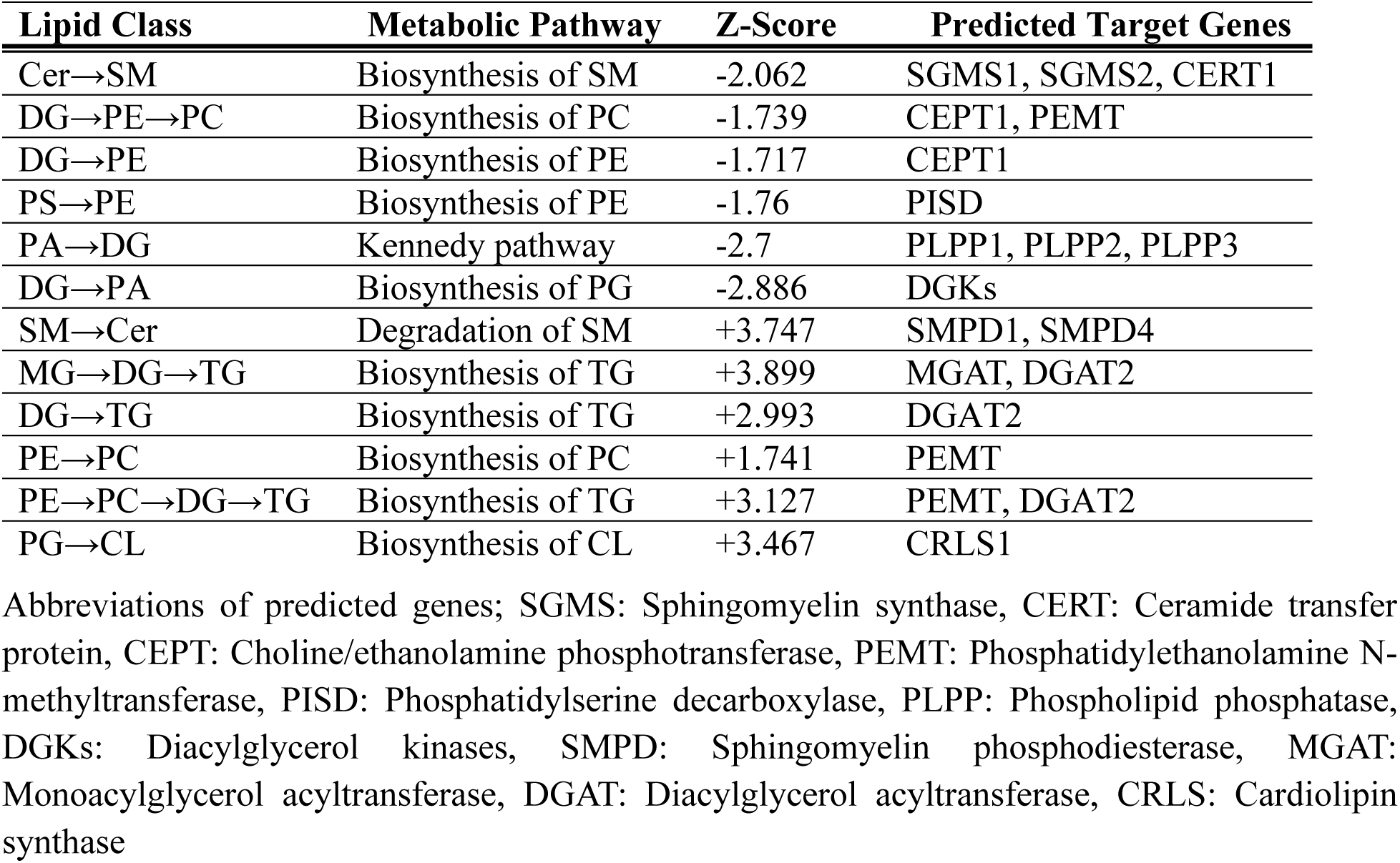
DMPA driven changes in lipid metabolic pathways in the vaginal epithelial barrier. BioPAN based pathway mapping predicted metabolic shifts across three major lipid classes: glycerolipids, phospholipids, and sphingolipids along with their respective predicted genes for the enzymes driving the reaction. Negative and positive Z-score values indicate significantly suppressed and enhanced pathways, respectively.

## 4. DISCUSSION

In this discovery lipidomics study, we quantitatively profiled the lipidomic landscape of the female genital tract in high-risk, HIV-negative women sex workers and identified markedly altered lipid signatures associated with exposure to injectable DMPA and oral hormonal contraceptive pills (OCPs). The identified lipids belong to 15 subclasses including several structural and signaling phospholipids, sphingolipids, energy/storage lipids, and sterols (Fig. 2A).

The CVLs from women using progestin-only injectable DMPA and the estrogen containing combined OCP exhibited consistent divergent effects across the cervicovaginal lipidome abundance. Our result demonstrated marked downregulation of structural and signaling membrane lipid subclasses such as phospholipids (PC, Lyso PC, PA, PE) sphingolipids (ceramide, sphingomyelin, glycosphingolipids), and upregulation of storage lipids (triglycerides) and sterols in the injectable DMPA group when compared to OCP and no hormone (NH) groups (Fig. 3-5). Among the most depleted phospholipids in the CVL of DMPA users were several phosphatidylcholine species such as PC(P-16:0/15:1), PC(15:0/18:1), PC(22:1/20:0), PC(O-18:0/O-3:1), PC(P-18:1/22:6), PC(18:0/22:1), PC(20:1/13:0), PC(18:3/18:3), PC(17:2/12:0), and PC(18:1/18:2) (Fig 3C). With four exceptions, all the differentially affected phosphatidylethanolamine species including PE(14:1/22:2), PE(14:0/20:3), PE(17:1/17:1), PE(O-20:0/18:0), Lyso PE(0:0/22:6), PE(20:4)/15:1), PE(21:0/20:0), PE(20:1/12:0), PE(O-16:0/13:0), and PE(22:1/21:0) also showed significant downregulation in the DMPA users (Fig. S1A). Even if the change in abundance at subclass level of other phospholipids was non-significant, specific lipids including 4 PS, 5 PG, 1 CL, 4 PA, and 4 PI species exhibited significant reduction, while only 2 PA and 1 PI showed significant upregulation associated with DMPA (Fig. 3D). The most abundant membrane phospholipids in host mucosal epithelial cells, are PC and PE, and in microbial cells are PE and PG, ^27,28^. The dysregulated depletion of a wide range of host and microbial membrane phospholipids seen in our analysis could be a potential molecular mechanism for the well-studied off-target effects of DMPA including vaginal microbial dysbiosis, genital barrier integrity impairment, thinning and atrophy of the vaginal epithelium, and thereby increased susceptibility to HIV infection ^3,6,29–31^.

Another important membrane lipid class? significantly lowered in the DMPA group compared to the OCP and NH groups are sphingolipids, including Cer, sphingomyelin SM, and GSL species (Fig. 4A-D). Such a drastic and unidirectional DMPA-induced quantitative shift over a wide-range of sphingolipids may compromise their crucial role in maintaining membrane structure and modulating multiple signaling pathways that determine host cell’s fate and risk to infection ^32,33^. Notably, the observed dysregulation of ceramides in the vaginal tract of women using DMPA may reflect functional decline in regulating membrane integrity, inflammation, microbial invasion, cell death, proliferation, and autophagy in the mucosal epithelia ^34,35^. Metabolically, ceramide is a central biosynthetic precursor of other complex sphingolipids, and hence its aberrant down-regulation in the DMPA users would be predicted to result in decreased abundance of downstream products (SM and GSL), seen in our results (Fig. 4A-C). The decrease in SM and GSL abundances seen in the DMPA group can be associated with the impairment of their functions in tightly packing the membrane lipid rafts and downregulating immune cell activation signaling cascades, that may result in immunosuppression ^36,37^ and consequently increased susceptibility to genital HIV infection. Strikingly, the significant changes in galactosylceramide (GalCer) and glucosylceramide (GlcCer) glycolipids (Fig. 4B) are of interest as their expression on mucosal surfaces has recently been associated with modulation of host-pathogen interactions including HIV-1 infection ^38–40^, making them excellent targets for drug discovery. These results suggest that dysregulation of the lipids that maintain the cervicovaginal epithelial membrane and genital microbial composition are likely involved in the complex crosstalk among hormonal contraceptives, host, and microbial communities.

In contrast to the structural and signaling lipid classes, our finding that DMPA users showed marked elevation in several TGs in the CVLs (Fig. 5A, C, D) is in line with the stimulatory effect of progesterone in maternal and fetal hepatic lipogenesis ^41^, although its effect on TG metabolism in the vaginal tract has been hardly reported. Interestingly, consistent depletion in cervicovaginal phospholipids and sphingolipids, as well as increase in TG, as observed in the DMPA users, has been reported as prominent component of cervix shortening and preterm birth ^14,42^. Specifically, hyperlipidemia with elevated levels of triglyceride and cholesterol has also been shown to shape the FGT, leading to chronic inflammation, reproductive failure, dysregulated epithelial tight junctions, and increased pathogen susceptibility ^18,43^. However, whether the DMPA driven elevated genital TG is due to its systemic or local effect needs to be explored further.

In the women using OCP, the concomitant increase in genital FFAs and DG, along with the reduction in their metabolic product (TG) (Fig. 6A-D), implies that likely estrogen drives genital lipid metabolism reprogramming very differently than DMPA. The OCP-induced upregulation of FFAs may explain the protective effect of estrogen hormone in the FGT as FFAs can selectively enhance the growth of beneficial (eubiotic) vaginal microbes that protect against pathogens and improve vaginal health^44,45^. In a more detail analysis, microbially produced genital lipids differentially accounted for the hormonal contraceptive induced lipidomic shifting (Fig. 7), supporting the divergent effects of DMPA and OCP on genital lipid metabolism. These findings suggest an untapped interplay among sex hormones, lipidome, and microbiome in the FGT. Differences in molecular structures of lipids such as type of head group, fatty acid chain length, degree of unsaturation, cis-trans isomerism, and iso-branching are characteristic features that determine the complexity and functional diversity of lipid species ^19,46^. Both the host and the microbial community can synthesize even carbon fatty acid containing lipids, whereas odd carbon lipids are pre-dominantly synthesized by microbial communities, conferring dissimilarity in their recognition by host immune receptors ^20,26,47^. These distinctions in lipid biosynthesis and immune responses can be leveraged as promising avenue for host-directed or pathogen targeted-drug repurposing and development research.

Through the analysis of differential metabolic pathways, we found that DMPA suppressed the de novo phospholipid and sphingolipid biosynthesis pathways, further supporting our findings of aberrantly altered membrane and signaling lipids in FGT. On the other hand, this injectable contraceptive appears to enhance salvaging of various genital lipids from reaction intermediates and cell membrane lipid pool, shifting them towards TG accumulation (Fig. 8 and Table 1). For instance, the DMPA-driven concurrent depletion of PC, lyso PC, and FFAs indicates that the most abundant membrane phospholipid, PC, is being pooled to TG via DG by changing its head group, rather than its cleavage to lyso PC and FFA. This DMPA driven switching of membrane lipids to stored lipid droplets is mechanistically worth noting as it may lead to thinning of mucosal epithelial barrier, chronic inflammation, and increased susceptibility to HIV.

In conclusion, our comprehensive genital lipidomics study revealed an interesting connection between hormonal contraceptives and essential lipids that potentially impact vaginal microbiota, immune regulation, barrier integrity, and pathogen susceptibility. Our main findings of depleted membrane and signaling phospholipids, ceramides, sphingomyelins, glycosphingolipids, along with upregulated TG in the DMPA users provides important insights that may account for the DMPA associated increase in susceptibility to HIV infection in the FGT. These findings underscore the need to address cervicovaginal lipidomic dysregulation associated with HCs while designing interventions to improve women’s reproductive health. While this study provides substantial contributions to existing literature, functional studies that examine HC driven dysregulation of genital lipid metabolism and its impact on HIV susceptibility are needed to confirm these studies.

## Author contributions

CK, DW, and AGG conceived the metabolomics study. AGG, NH, and YAA optimized analytical method. CK, JMW, JL and KRF designed and got approvals for the original study. JL, JK, KRF, provided on site project supervision, collected, and provided CVL samples. CK provided project supervision and acquired project funding. AGG and MN performed all lipidomics experiments, data processing, analysis, and visualization. AGG wrote the original draft. YAA, NH, DW, and CK reviewed and edited the manuscript. All authors reviewed and approved the manuscript.

## Acknowledgements

The authors would like to sincerely thank all the women who provided samples that were analyzed in this study. We also acknowledge staff of the Center of Microbial Chemical Biology (CMCB) at McMaster University for their technical assistance.

## Data availability

All data related to this study can be made available upon request.

