## Supplementary Materials for "Hormonal Contraceptives Drive Genital Lipid Metabolism Reprogramming and Susceptibility to HIV Infection"

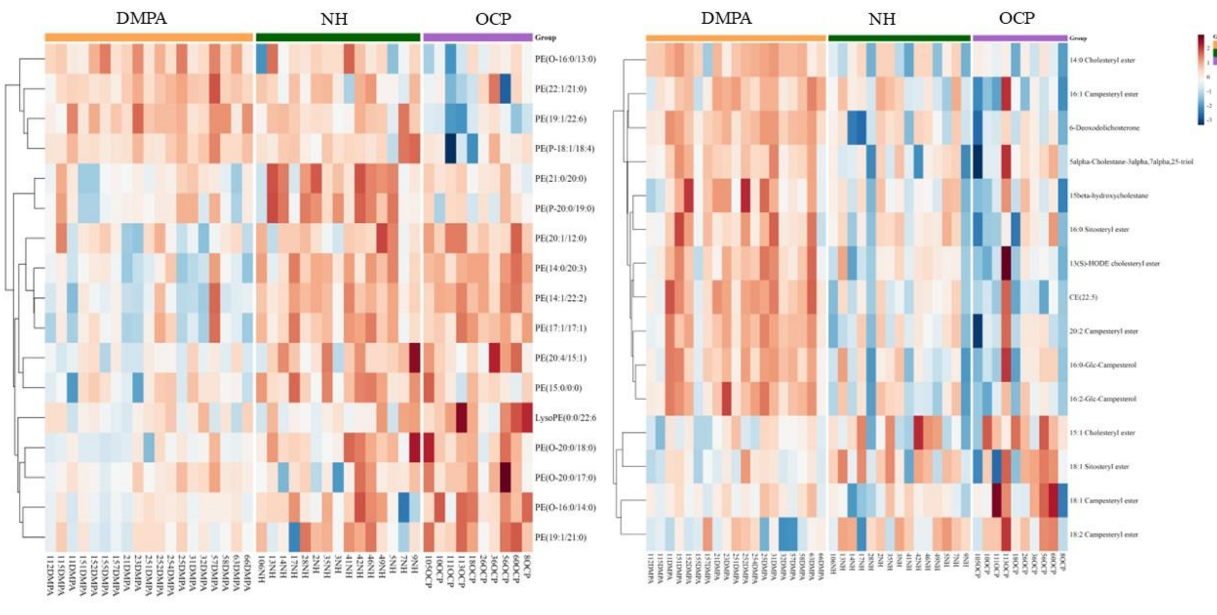


A) Phosphatidylethanolamine (PE)

B) Sterols

**Fig. S1**: Differentially altered PE (**A**) and sterol (**B**) lipid species among DMPA, NH, and OCP taking groups. Rows represent individual lipid species and columns correspond to study groups: depot medroxyprogesterone acetate (DMPA), no hormone (NH), and oral contraceptive pill users (OCP).

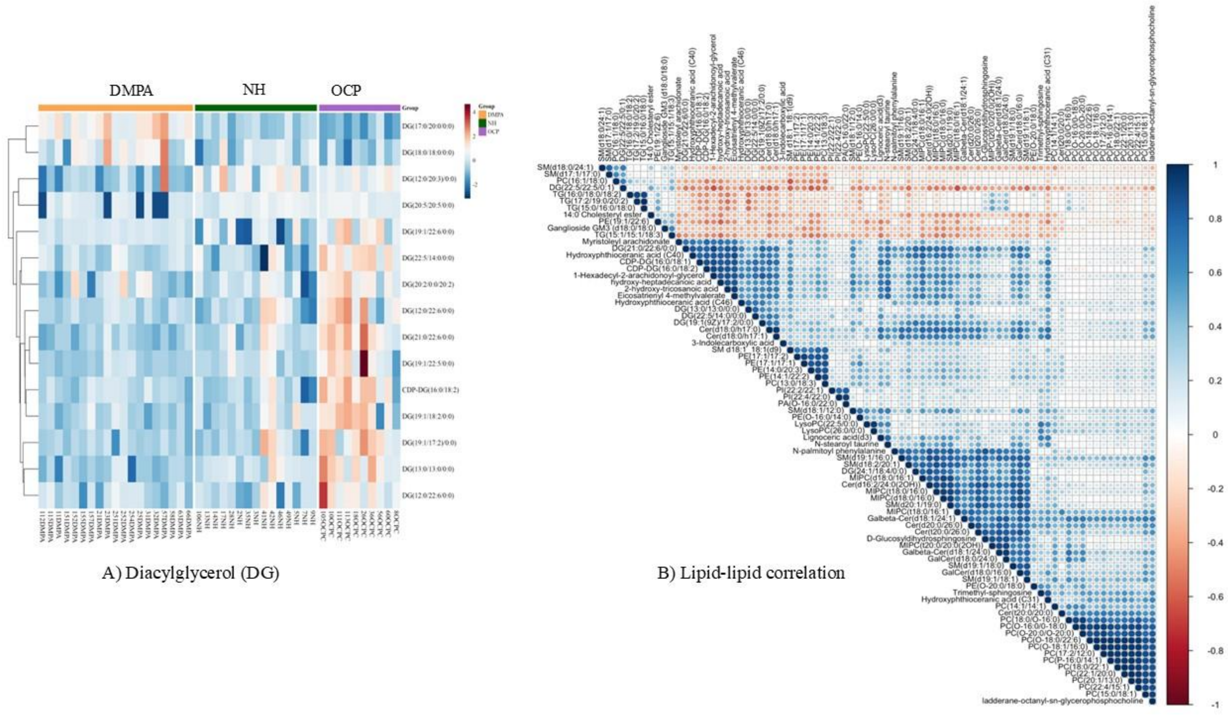


A) Diacylglycerol (DG)

B) Lipid-lipid correlation

**Fig. S2**: Differential and correlation pattern of genital lipids. **A**) OCP associated upregulation of specific DG lipids in the OCP group. **B**) correlation heatmap showing pairwise lipid-lipid co-regulation among 90 lipid species identified as significant in the differential analysis.

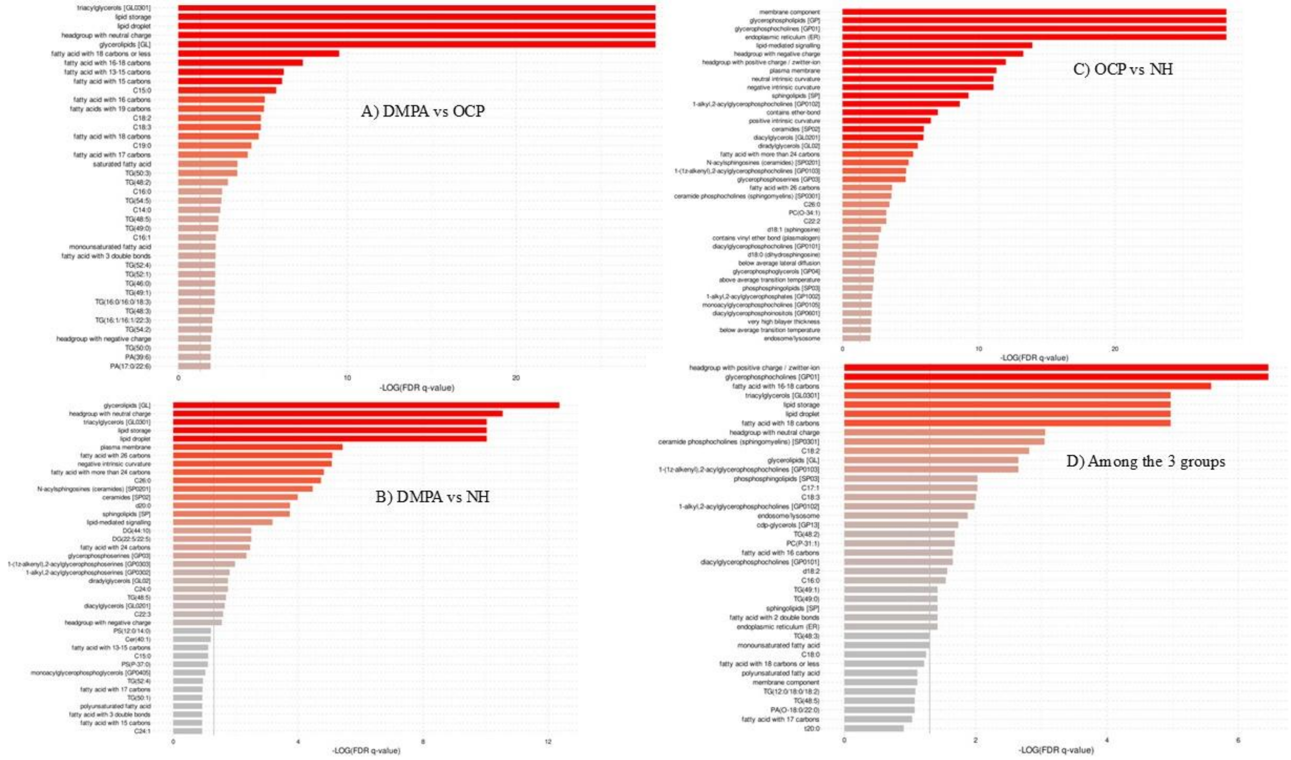


**Fig. S3**: **Lipid Ontology (LION) enrichment analysis of genital lipidomes.** Bar charts representing LION-term enrichment in ranking mode for pairwise comparisons: (**A**) DMPA vs. OCP, (**B**) DMPA vs. NH, and (**C**) OCP vs. NH, **D**) Among all groups. Y-axes list enriched LION-terms categorized by chemical class, biophysical property, or cellular location. X-axes represent significance as $log_{10}$ (FDR *q-value*); the vertical dashed line indicates the significance threshold (*q> 0.05*). Bar colors are scaled by enrichment significance (grey to dark red).
